# Mental health service activity during COVID-19 lockdown among individuals with potential neurodevelopmental disorders: South London and Maudsley data on services and mortality from January to July 2020

**DOI:** 10.1101/2020.10.11.20210625

**Authors:** Evangelia Martin, Eleanor Nuzum, Matthew Broadbent, Robert Stewart

## Abstract

The lockdown and social distancing policy imposed due to the COVID-19 pandemic is likely to have had a widespread impact on mental healthcare service provision and use. Previous reports from the South London and Maudsley NHS Trust (SLaM; a large mental health service provider for 1.2m residents in South London) highlighted a shift to virtual contacts among those accessing community mental health and home treatment teams and an increase in deaths over the pandemic’s first wave. However, there is a need to quantify this for individuals with particular vulnerabilities, including those with learning disabilities and other neurodevelopmental disorders. Taking advantage of the Clinical Record Interactive Search (CRIS) data resource with 24-hourly updates of electronic mental health records data, this paper describes daily caseloads and contact numbers (face-to-face and virtual) for individuals with potential neurodevelopmental disorders across community, specialist, crisis and inpatient services. The report focussed on the period 1^st^ January to 31^st^ July 2020. We also report on daily accepted and discharged trust referrals, total trust caseloads and daily inpatient admissions and discharges for individuals with potential neurodevelopmental disorders. In addition, daily deaths are described for all current and previous SLaM service users with potential neurodevelopmental disorders over this period. In summary, comparing periods before and after 16^th^ March 2020 there was a shift from face-to-face contacts to virtual contacts across all teams. The largest declines in caseloads and total contacts were seen in Home Treatment Team, Liaison/A&E and Older Adult teams. Reduced accepted referrals and inpatient admissions were observed and there was an 103% increase in average daily deaths in the period after 16^th^ March, compared to the period 1^st^ January to 15^th^ March (or a 282% increase if the 2-month period from 16^th^ March to 15^th^ May was considered alone).

## Background

The COVID-19 pandemic has presented unique challenges for mental healthcare provision due to a combination of factors, including the impact of the virus itself, the impact of social distancing policies on delivery of mental healthcare, as well as potential psychological effects of social distancing restrictions. Mental healthcare challenges have included the increased vulnerability of its patients, already-reduced life expectancies, pre-existing problems in healthcare access, increased staff sickness and isolation, COVID-19 infections (suspected or confirmed) across inpatient and outpatient services, and the need to minimize face-to-face contacts (1). Impacts may be higher for people with learning disabilities and other neurodevelopmental disorders due to changes in healthcare provision and increased physical and mental health multimorbidity. However, these have received little description to date.

We have previously reported on the mental healthcare impact of the UK COVID-19 pandemic via the Clinical Record Interactive Search (CRIS) data platform which receives 24-hourly updates from its source electronic mental health records. Previous pandemic era reports are listed on https://www.maudsleybrc.nihr.ac.uk/facilities/clinical-record-interactive-search-cris/covid-19-publications/. This has included a sizeable shift from face-to-face to virtual contacts in community mental health teams and home treatment teams, as well as a decrease in caseloads and total contacts in home treatment teams (2). We are seeking to extend this analysis to groups who might have specific needs and service use characteristics that might not be well reflected in Trust-wide data. This report focuses on service users with potential neurodevelopmental disorders.

## Methods

The Biomedical Research Centre (BRC) Case Register at the South London and Maudsley NHS Foundation Trust (SLaM) has been described previously (3;4). SLaM serves a geographic catchment of four south London boroughs (Croydon, Lambeth, Lewisham, Southwark) with a population of around 1.2 million residents and has used a fully electronic health record (EHR) across all its services since 2006. SLaM’s BRC Case Register was set up in 2008, providing researcher access to de-identified data from SLaM’s EHR via the Clinical Record Interactive Search (CRIS) platform and within a robust security model and governance framework (5). CRIS has been extensively developed over the last 10 years with a range of external data linkages and natural language processing resources (6). CRIS is updated from SLaM’s EHR every 24 hours and thus provides relatively ‘real-time’ data. SLaM’s EHR is itself immediately updated every time an entry is made, which include date-stamped fields indicating patient contacts (‘events’) and those indicating acceptance of a referral or a discharge from a given service (or SLaM care more generally). Mortality in the complete EHR (i.e. all SLaM patients with records, past or present) is ascertained weekly through automated checks of National Health Service (NHS) numbers (a unique identifier used in all UK health services) against a national spine; this provides confirmation of date of death but no further information (e.g. on recorded causes). CRIS has received approval as a data source for secondary analyses (Oxford Research Ethics Committee C, reference 18/SC/0372) and has supported over 200 peer reviewed publications to date

Activity and caseload data were extracted via CRIS and enumerated for every day from 1^st^ January 2020 to 31^st^ July 2020. The focus of this report is on clinical activity and mortality of individuals diagnosed with potential neurodevelopmental disorders; this was defined on the basis of ICD-10 codes F7x, F84x or F90x within structured fields for primary or secondary diagnoses in the source electronic health record (mandatory fields for all active caseloads), supplemented by ‘leaning dis*’ applied to output from a bespoke natural language processing algorithm developed within CRIS to extract text associated with diagnostic statements recorded in open text fields (https://www.maudsleybrc.nihr.ac.uk/facilities/clinical-record-interactive-search-cris/cris-natural-language-processing/). This supplementary approach allowed for the fact that neurodevelopmental disorders may be under-reported in coded diagnostic fields in favour of other co-occurring diagnoses.

Clinical activity was measured across 6 service areas: working age community Adult Mental Health Services (AMHS), Child and Adolescent Mental Health Services (CAMHS), Mental Health of Older Adults (MHOA), Early Intervention in Psychosis (EIP), Home Treatment Team and crisis services (HTT), and Liaison Accident & Emergency (A&E) services. Total Trust caseloads, number of accepted and discharged Trust referrals, number of inpatient admissions and discharges, and number of inpatients (total and those specifically under a Mental Health Act order) were also extracted for the client group. For AMHS, CAMHS, MHOA, EIP, HTT, Liaison, daily caseloads were calculated by ascertaining patients who were receiving active care from the service on a given day, based on the date a referral to that service was recorded as accepted to the point a discharge was made from that service. Daily contact numbers were ascertained from recorded ‘events’ (i.e. standard case note entries) for that service and were divided into the following groups according to structured compulsory meta-data fields for that event in the EHR: i) face-to-face contacts attended; ii) virtual contacts attended (by email, fax, mail, phone, online, or video link); iii) total contacts, as the sum of these two; iv) did not attend (DNA).

Finally, mortality data (number of deaths for all patients with potential neurodevelopmental disorders with SLaM records) were extracted for the period in question. Data extractions for this manuscript were carried out on 10^th^ September 2020.

Descriptive data were generated daily for the above parameters and displayed graphically. Weekend days and national holidays were omitted for AMHS, CAMHS, MHOA, and EIP contacts. For comparisons between time periods, 16^th^ March 2020 was chosen, as the date social distancing policy was announced by the UK government. Mean (SD) activity levels were described before and after this date and percentage changes quantified.

## Results

Mean daily statistics before and after 16^th^ March and proportional changes are displayed in Table 1. Weekly deaths in the period from 1^st^ January to 31^st^ July are displayed in Figure 1. Mean daily deaths for individuals with potential neurodevelopmental disorders significantly increased by 103% in the period after 16^th^ March compared to the period between 1^st^ January to 15^th^ March, although the primary excess was observed up to mid-May. If the 2-month period from 16^th^ March to 15^th^ May was taken as a comparison instead, the mean daily deaths rose to 0.97, a difference of 0.71 and an increase of 282%.

**Table 1.**
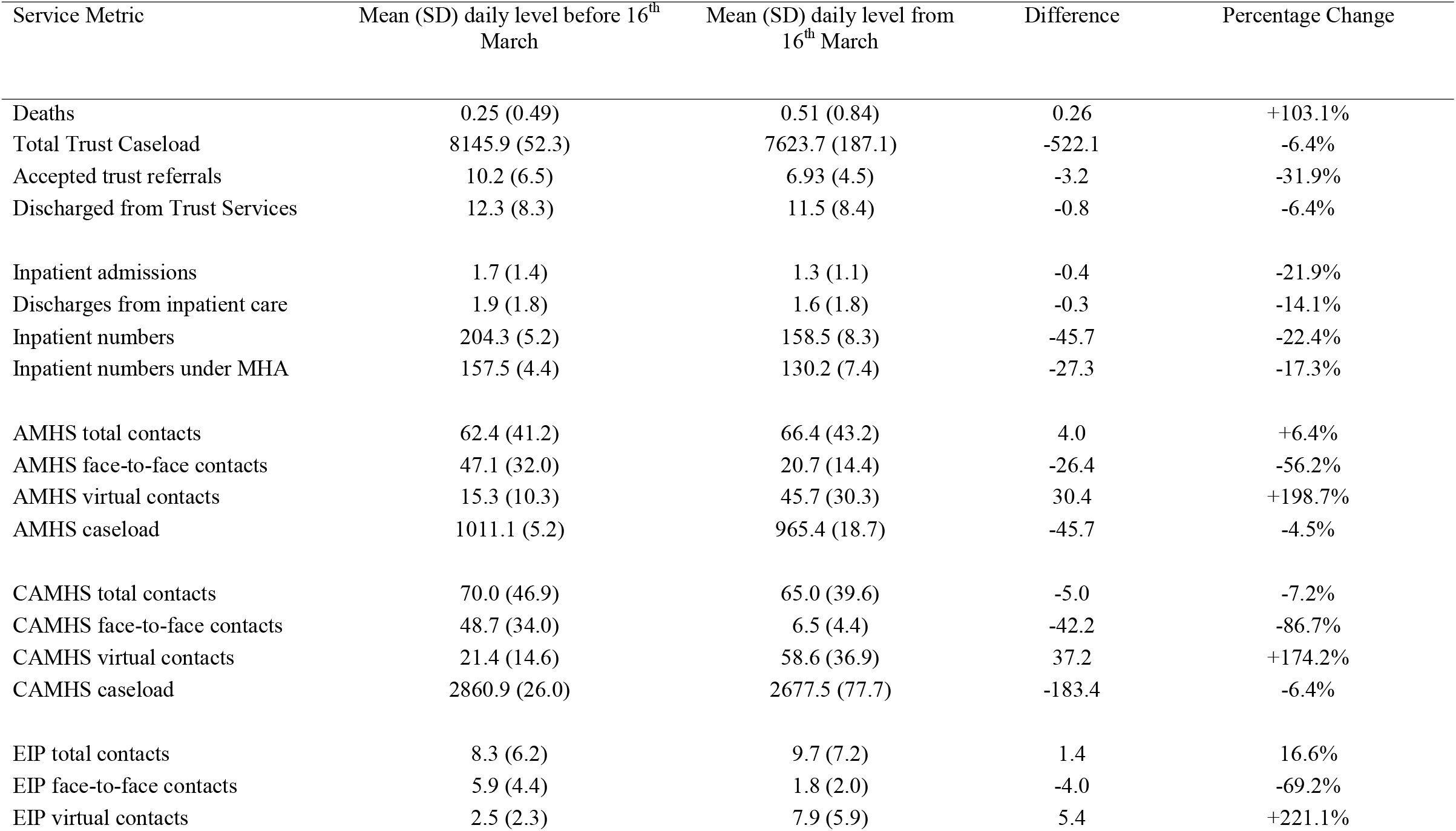
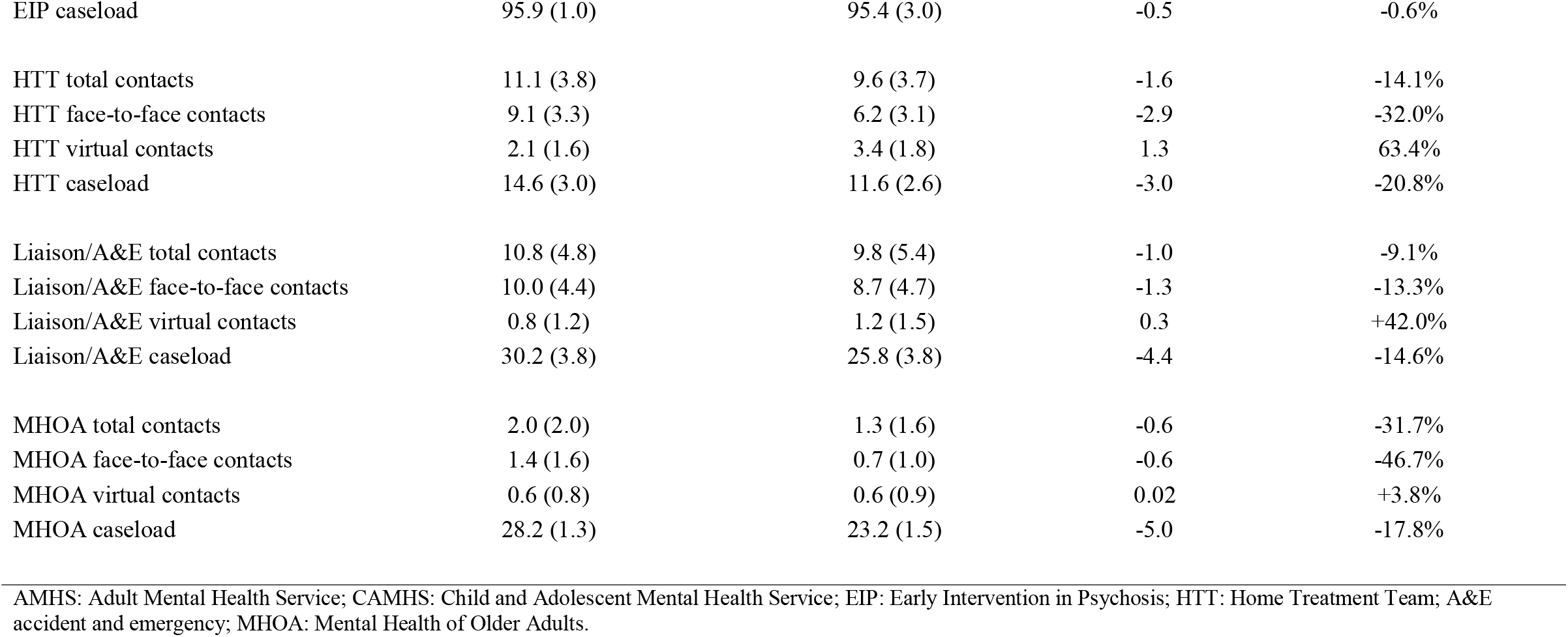
Levels and changes in daily service caseloads and contacts in patients with potential neurodevelopmental disorders from 1^st^ January to 31^st^ July 2020

**Figure 1:**
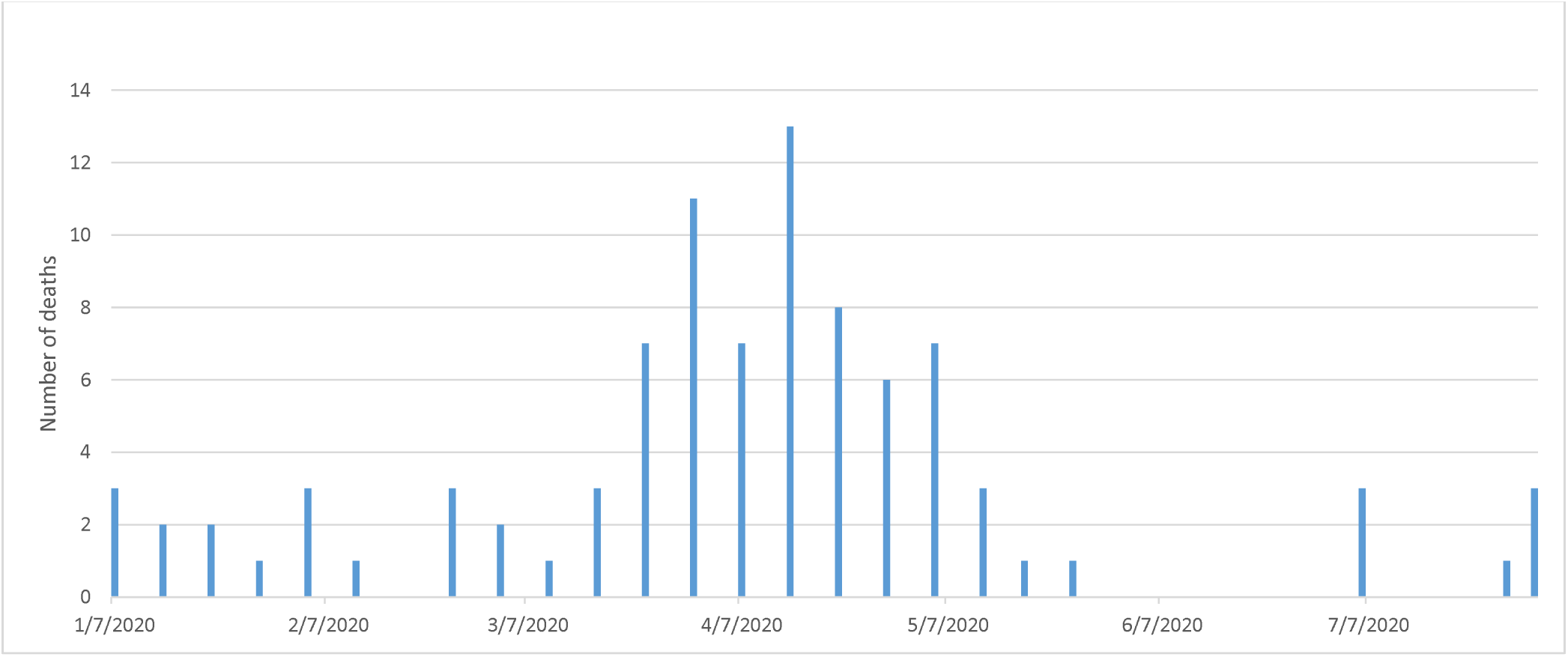
Total number of deaths (weekly; January-July 2020)

The daily Trust total caseload for service users with potential neurodevelopmental disorders is displayed in Figure 2, and daily accepted and discharged trust referral numbers are displayed in Figure 3. The mean number of accepted referrals decreased by 32%, and the number of discharges showed a relatively small reduction by 6%. The mean overall daily Trust caseload was reduced by 6%.

**Figure 2:**
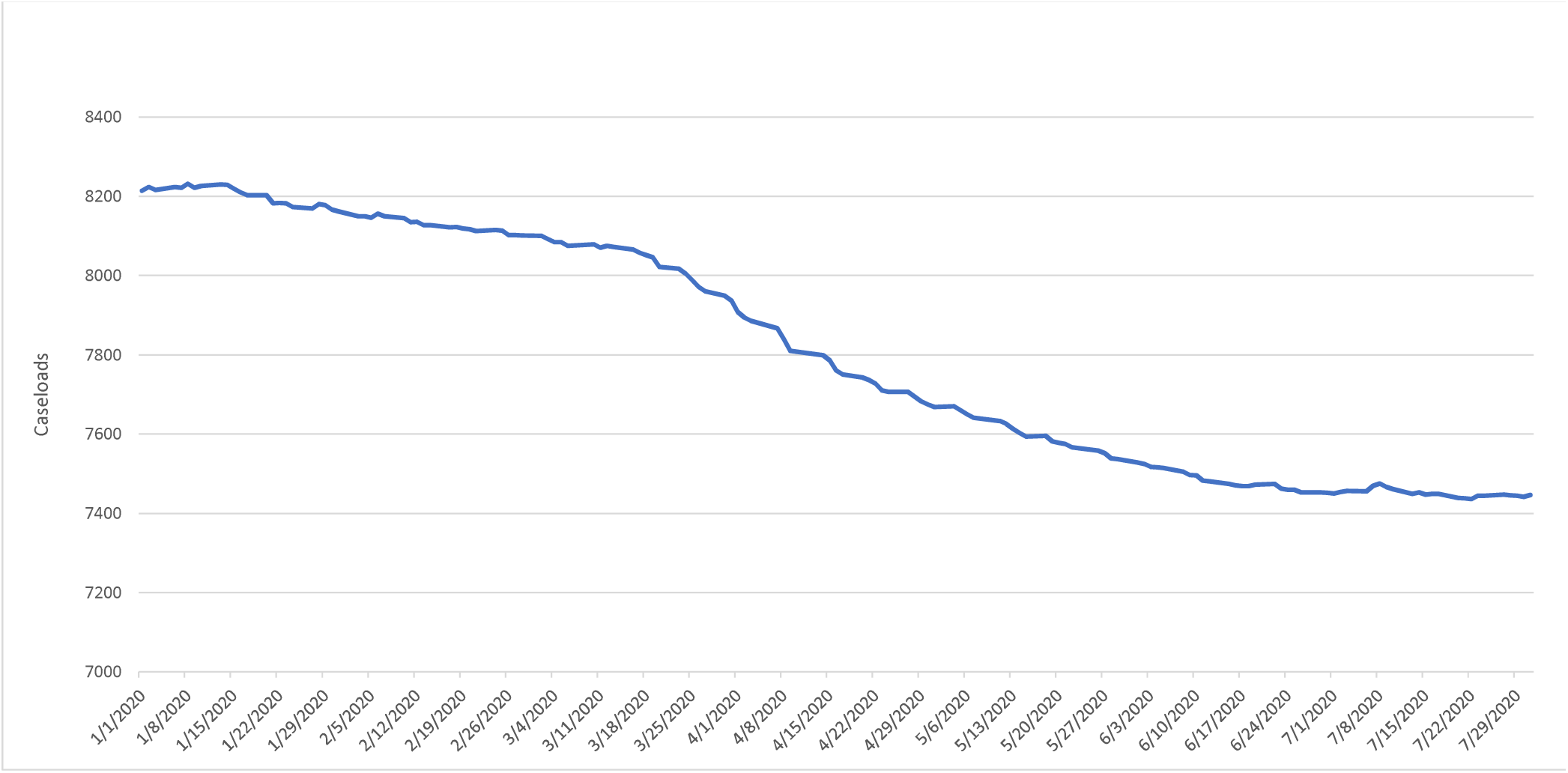
Trust total caseload (daily)

**Figure 3:**
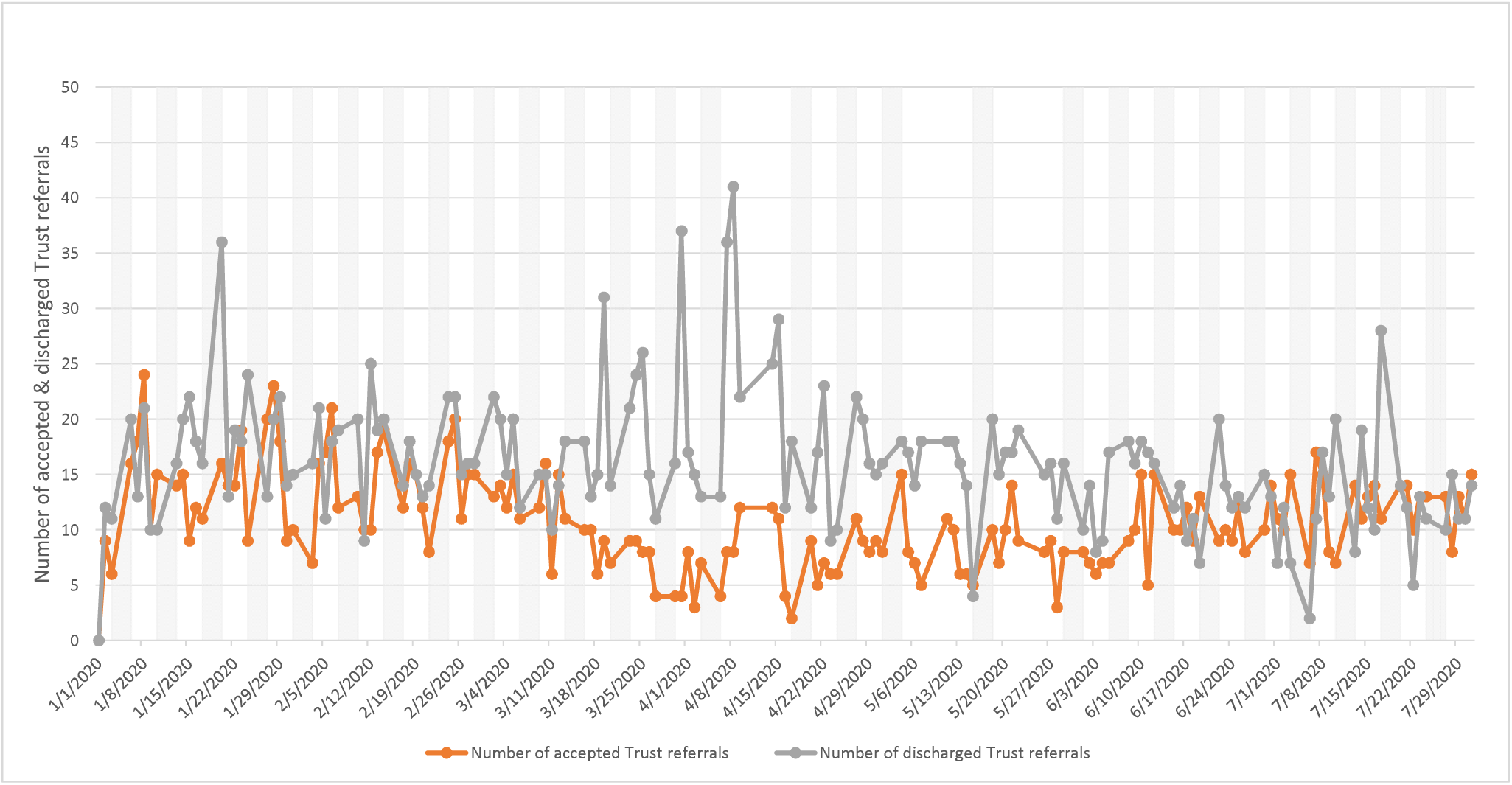
Number of accepted & discharged Trust referrals.

Data on inpatient care are displayed in Figures 4-5, showing a fall in daily inpatient numbers (by 22% overall and by 17% for those under the Mental Health Act), reflecting a high level of discharges in late March and early April (Figure 4).

**Figure 4:**
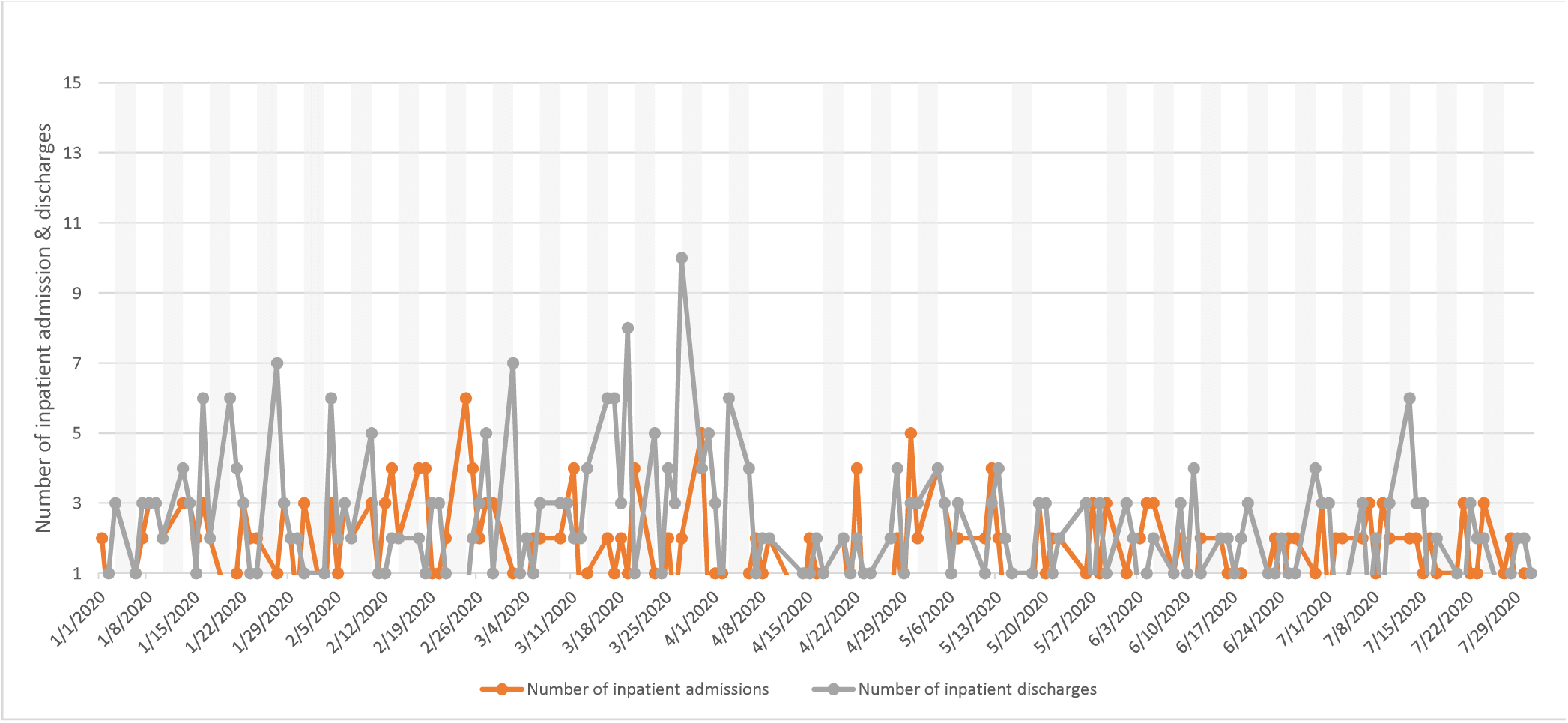
Number of inpatient admissions & discharges.

**Figure 5:**
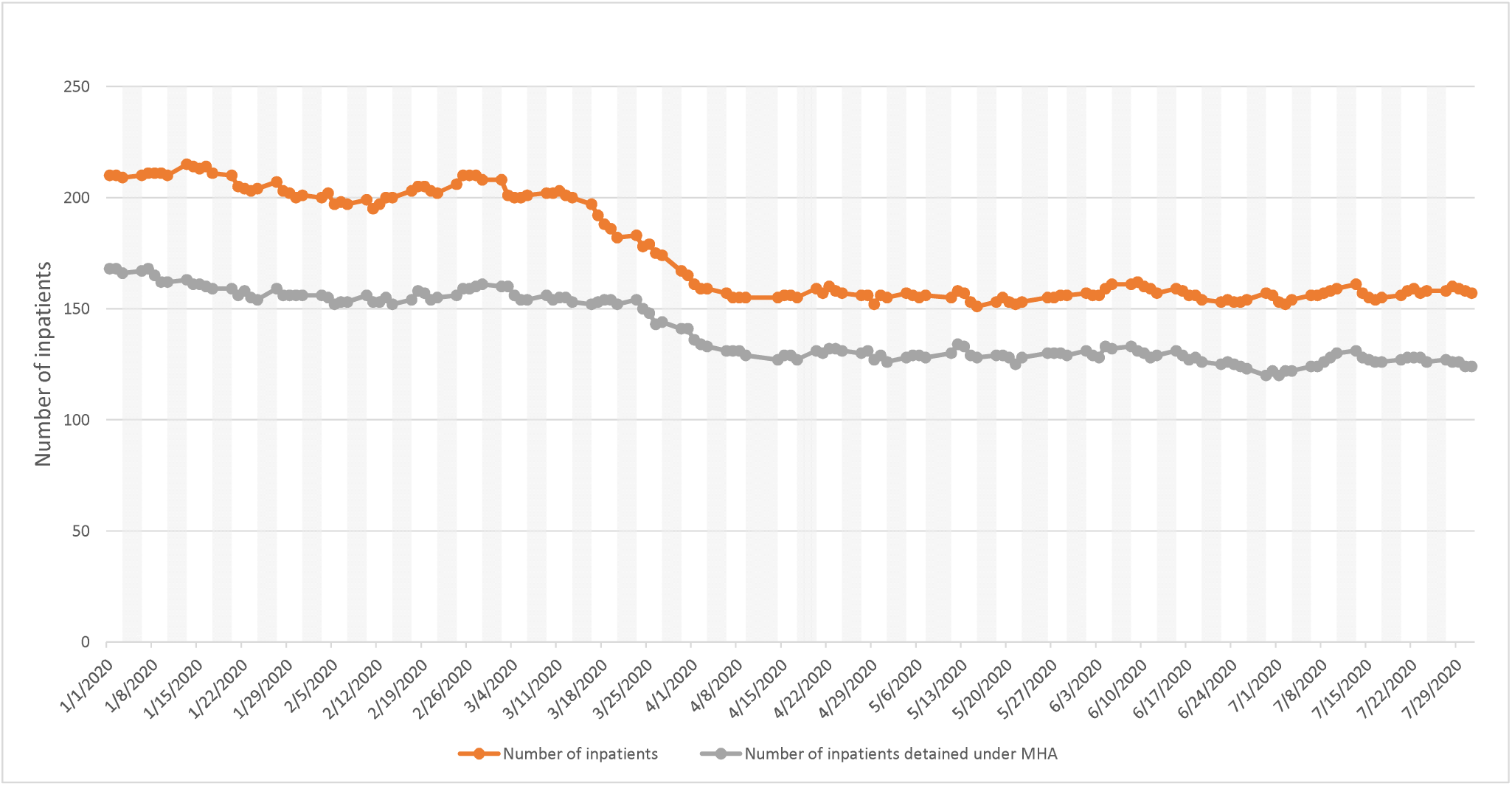
Number of inpatients / inpatients detained under the Mental Health Act.

AMHS daily contacts are displayed in Figure 6 and daily caseloads are displayed in Figure 7. Mean total daily contacts showed a relatively small increase (6%) overall after 15^th^ March. Face-to-face contacts were reduced by 56%, and virtual contacts were increased by 199%. However, mean daily caseloads were reduced only marginally by 5%.

**Figure 6:**
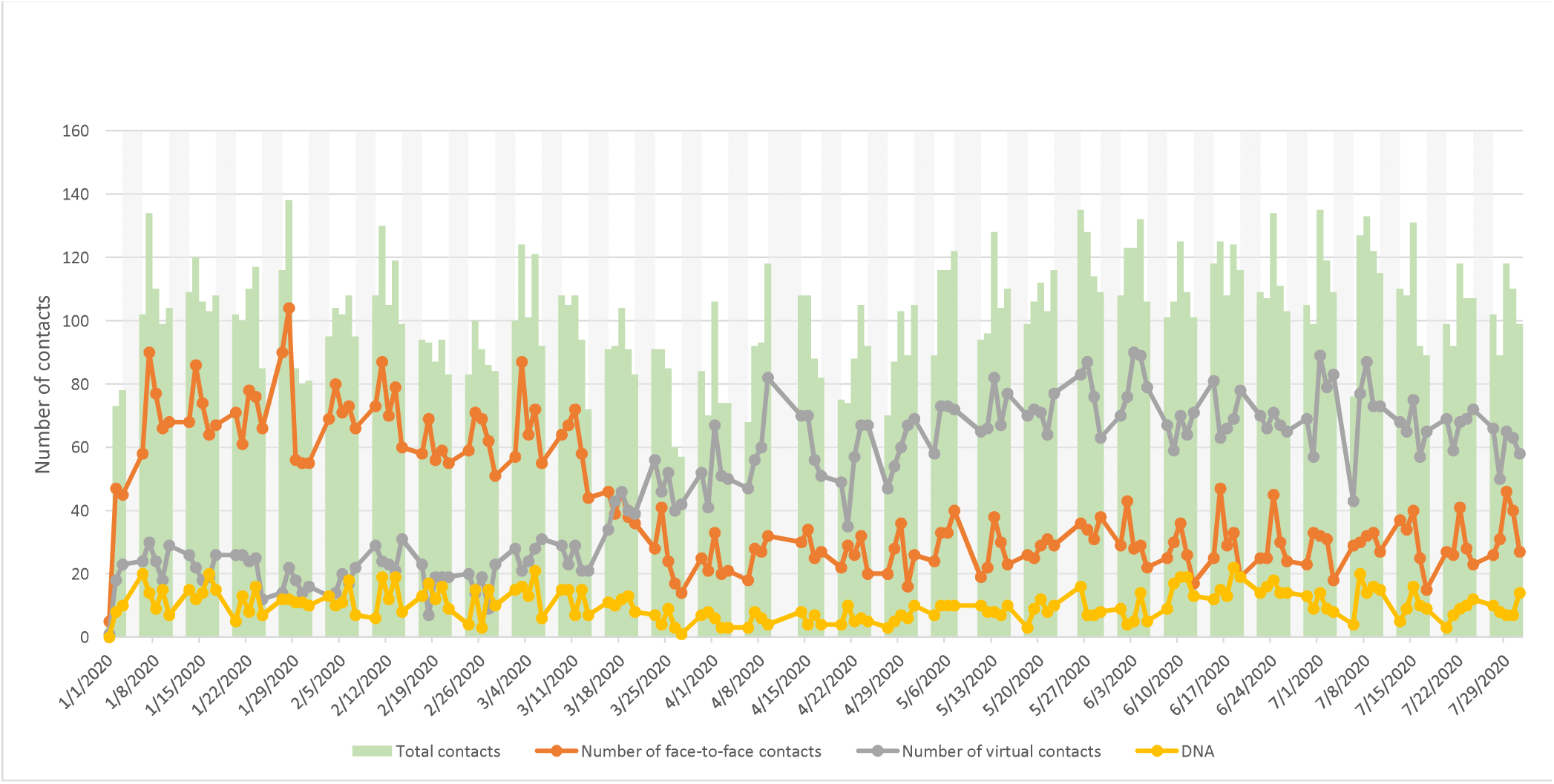
Adult Mental Health: non-specialist working age community services contacts by type (daily; January - July 2020)

**Figure 7:**
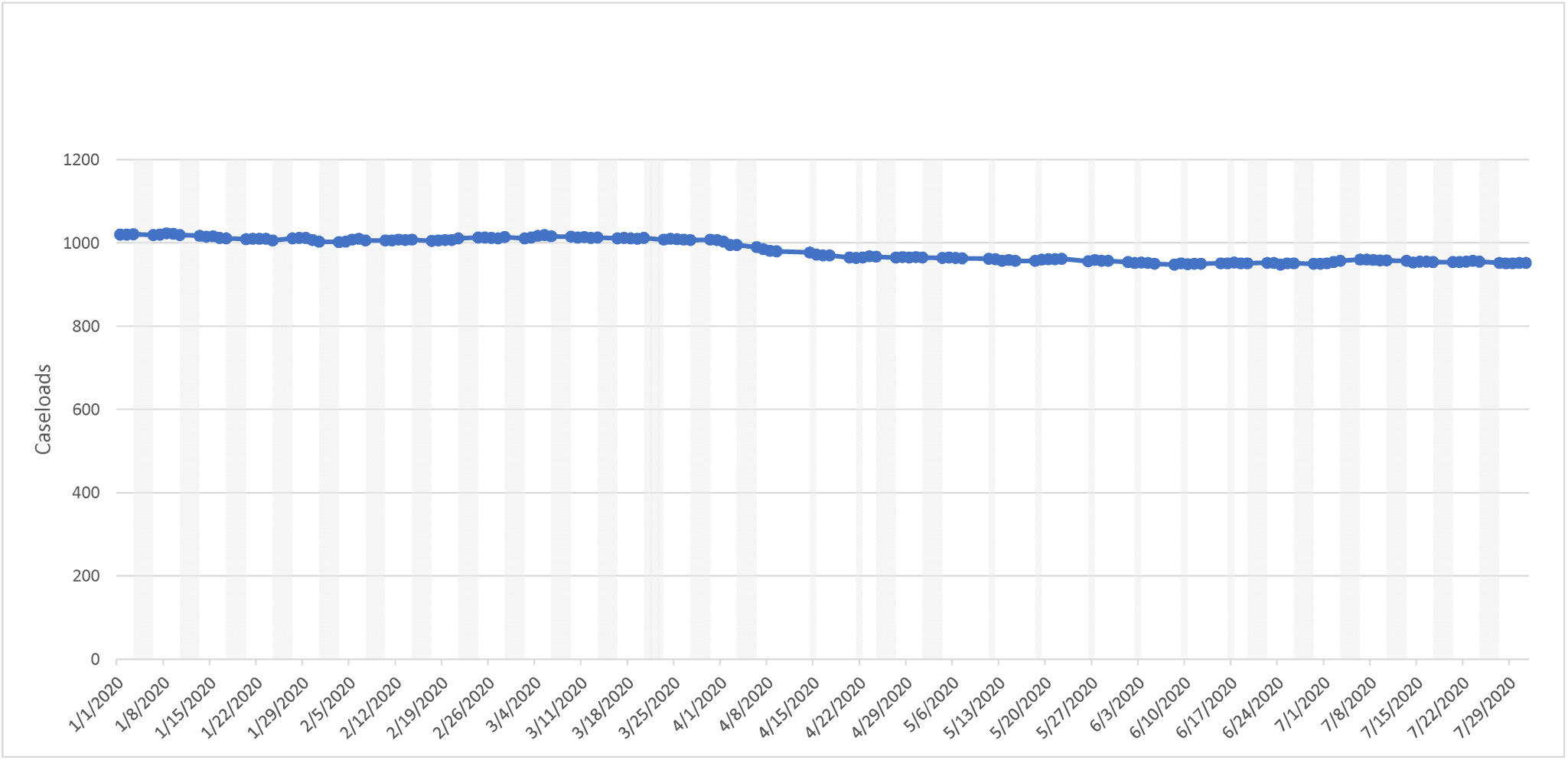
Adult Mental Health: non-specialist working age community services caseloads (daily; January - July 2020)

CAMHS daily contacts are displayed in Figure 8 and daily caseloads are displayed in Figure 9. Mean total daily contacts were reduced by 7%, with an 87% reduction in face-to-face contacts, and a 174% increase in virtual contacts. Mean daily caseloads were reduced by 6%.

**Figure 8:**
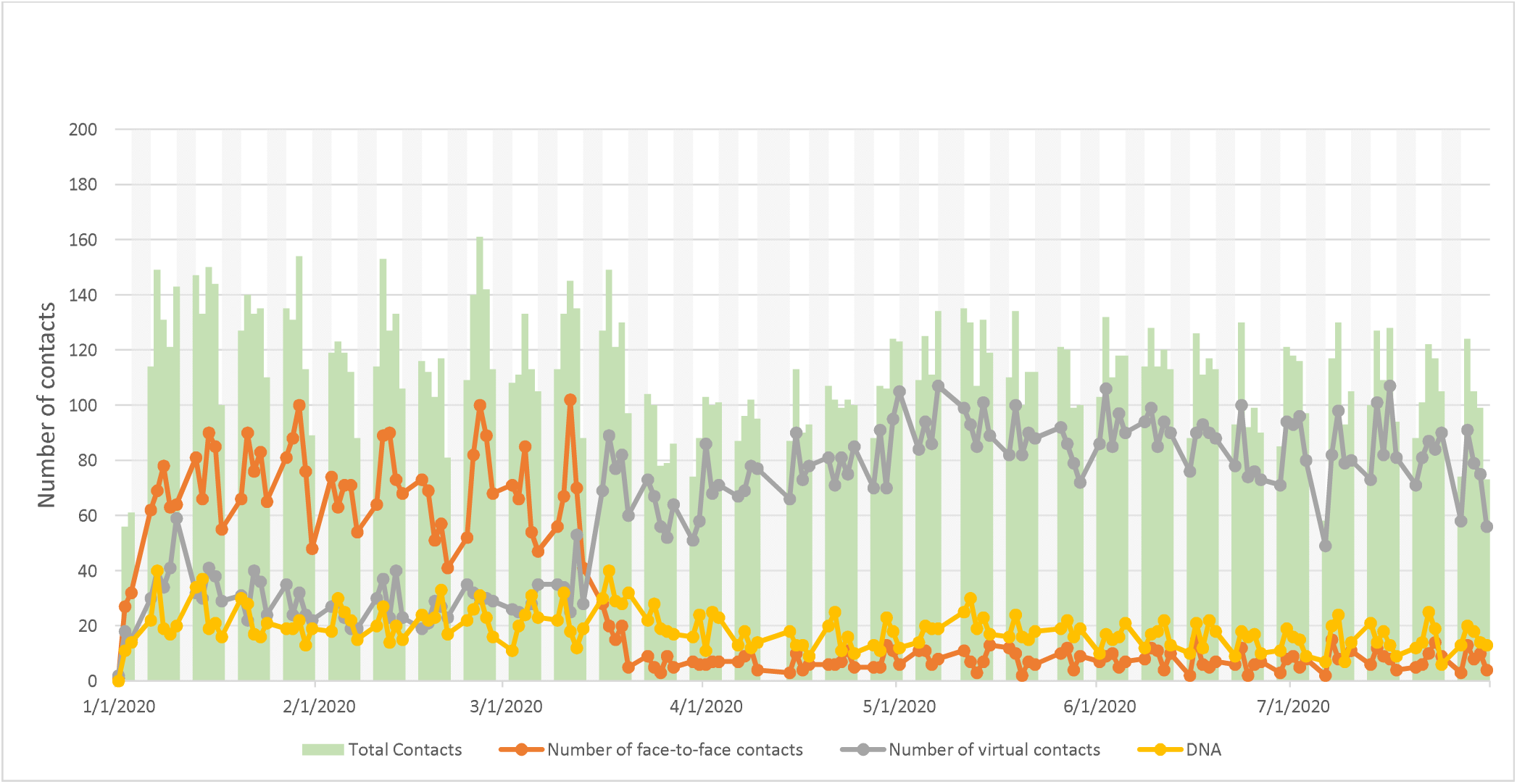
Child and Adolescent Mental Health Services contacts by type (daily; January - July 2020)

**Figure 9:**
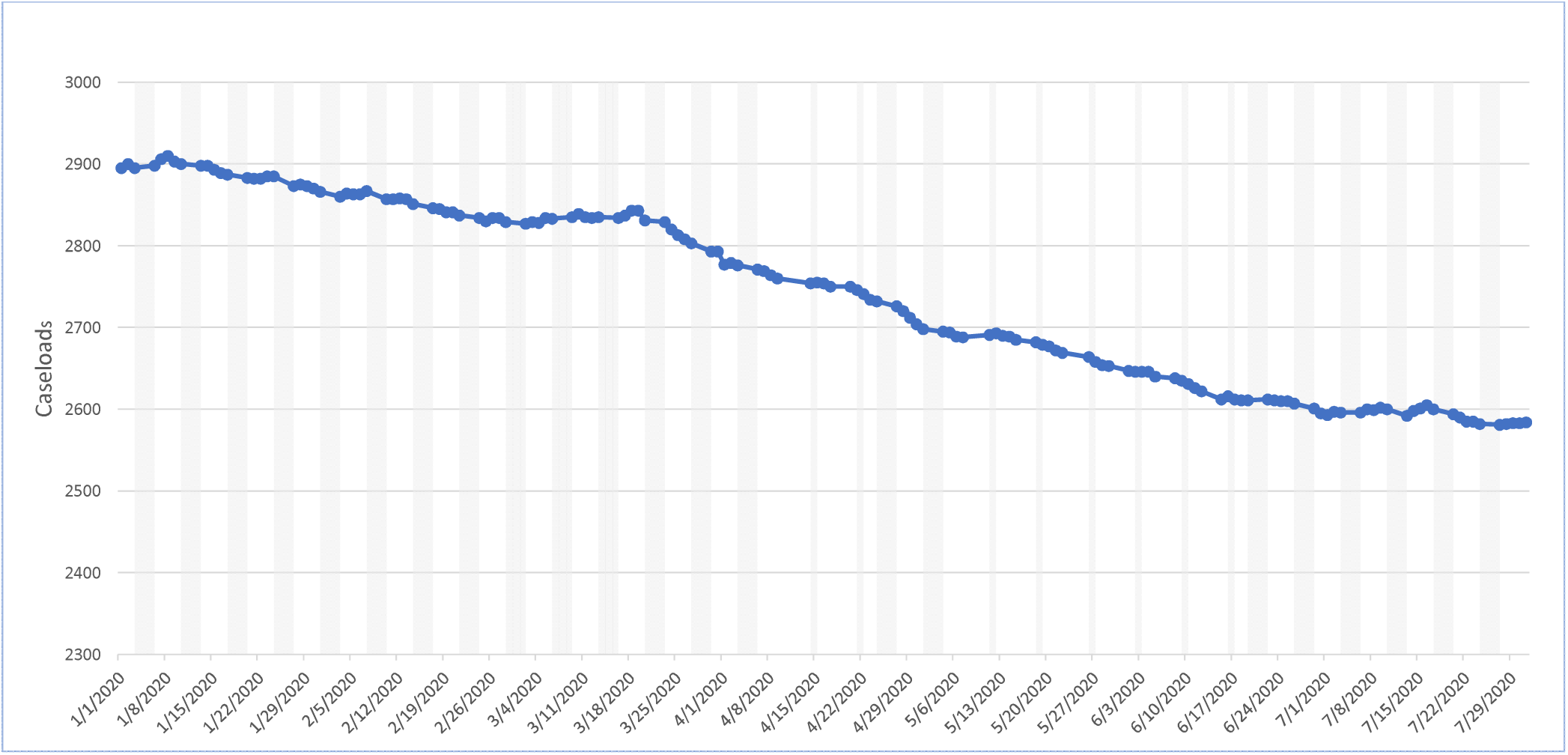
Child and Adolescent Mental Health Services caseloads (daily; January - July 2020)

EIP daily contacts are displayed in Figure 10 and daily caseloads are displayed in Figure 11. Mean total daily contacts showed an increase of 17%: a 69% reduction in face-to-face contacts accompanied by a 221% increase in virtual contacts. Mean daily caseloads remained relatively unchanged with a 1% increase.

**Figure 10:**
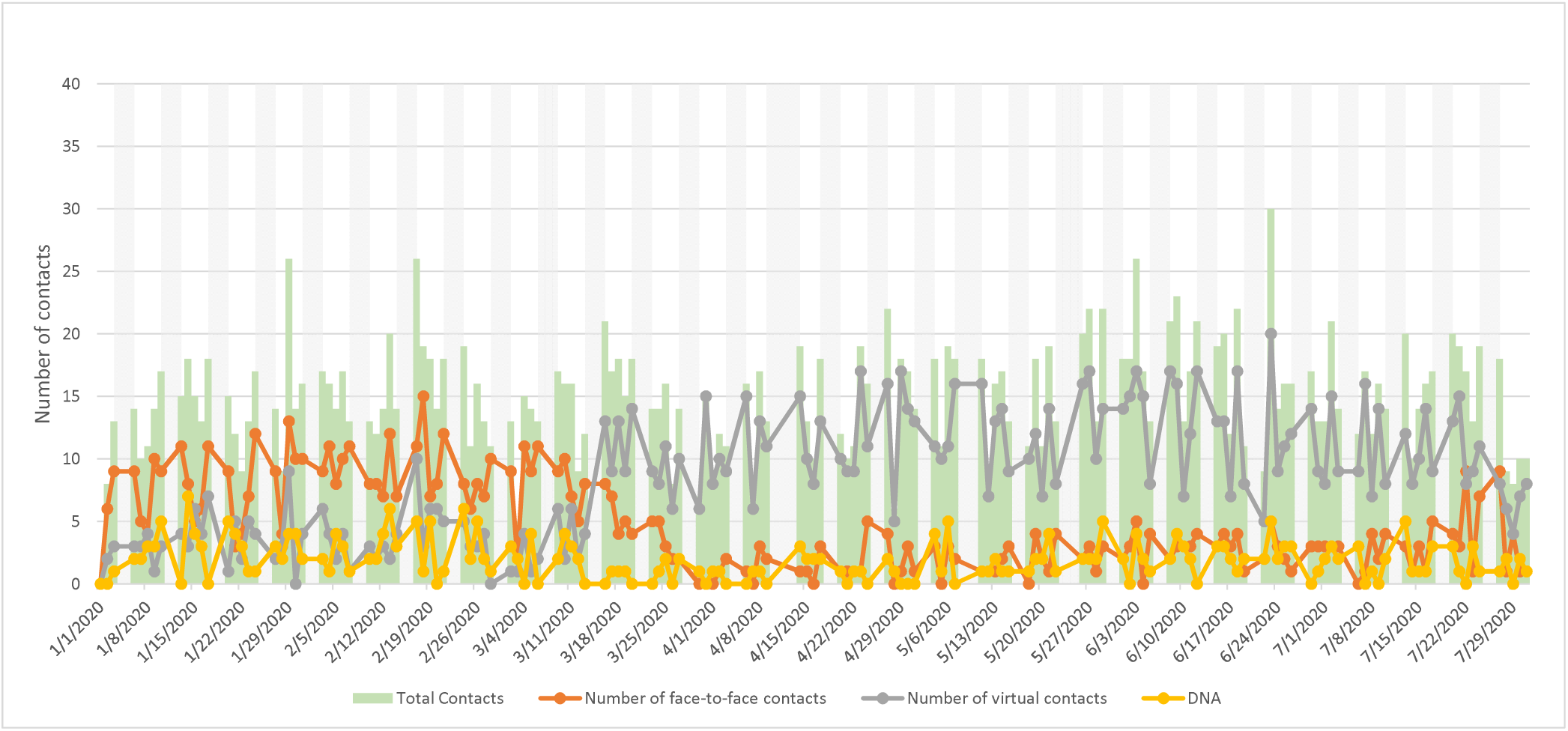
Early intervention psychosis services contacts per type (daily; January-July 2020)

**Figure 11:**
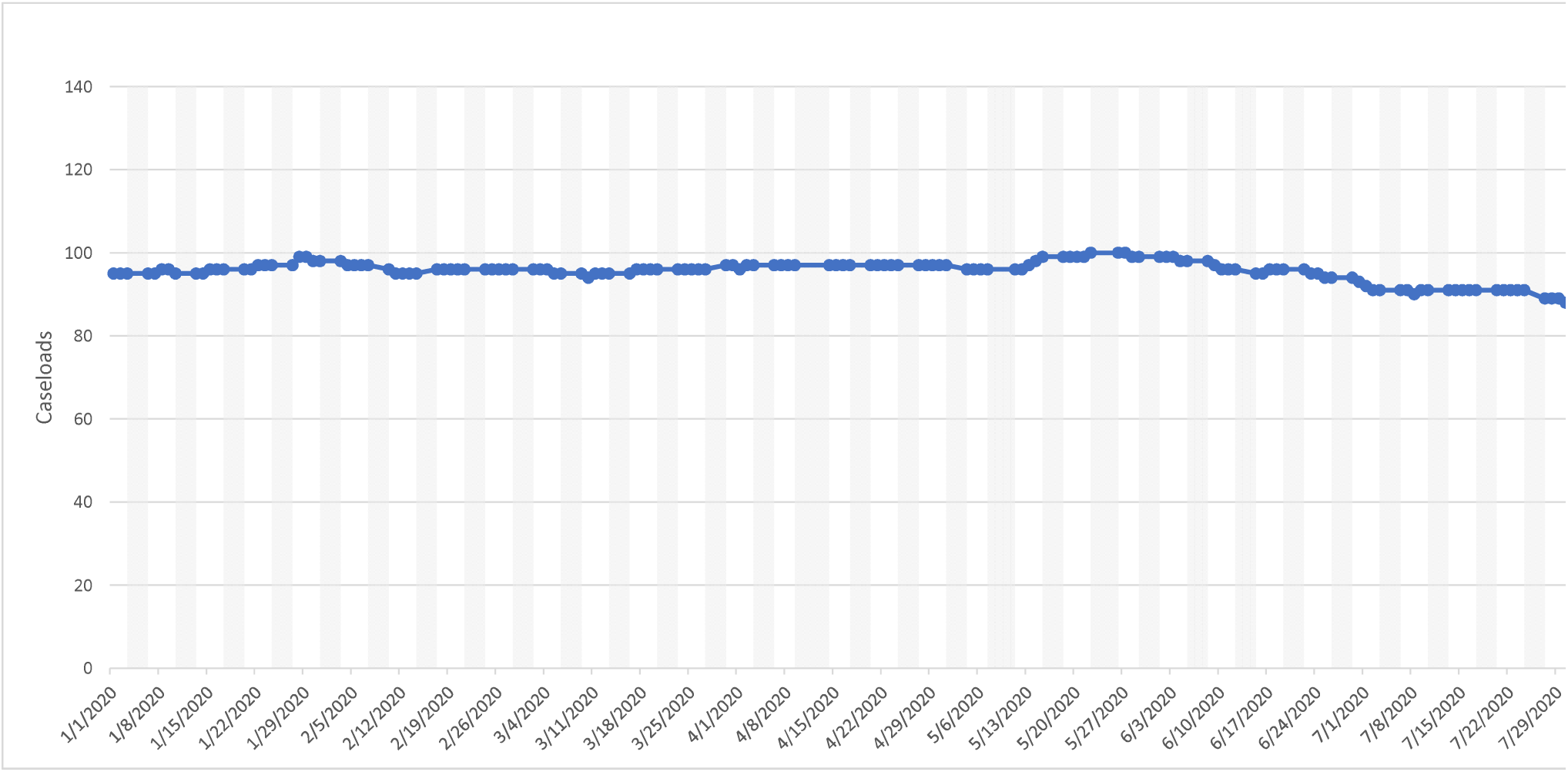
Early intervention psychosis services caseloads (daily; January-July 2020)

HTT daily contacts are displayed in Figure 12 and daily caseloads are displayed in Figure 13. Mean total daily contacts were reduced by 14% overall after 15^th^ March, made up of an 32% reduction in face-to-face contacts and a 63% reduction in virtual contacts. Mean daily caseloads were reduced by 21%.

**Figure 12:**
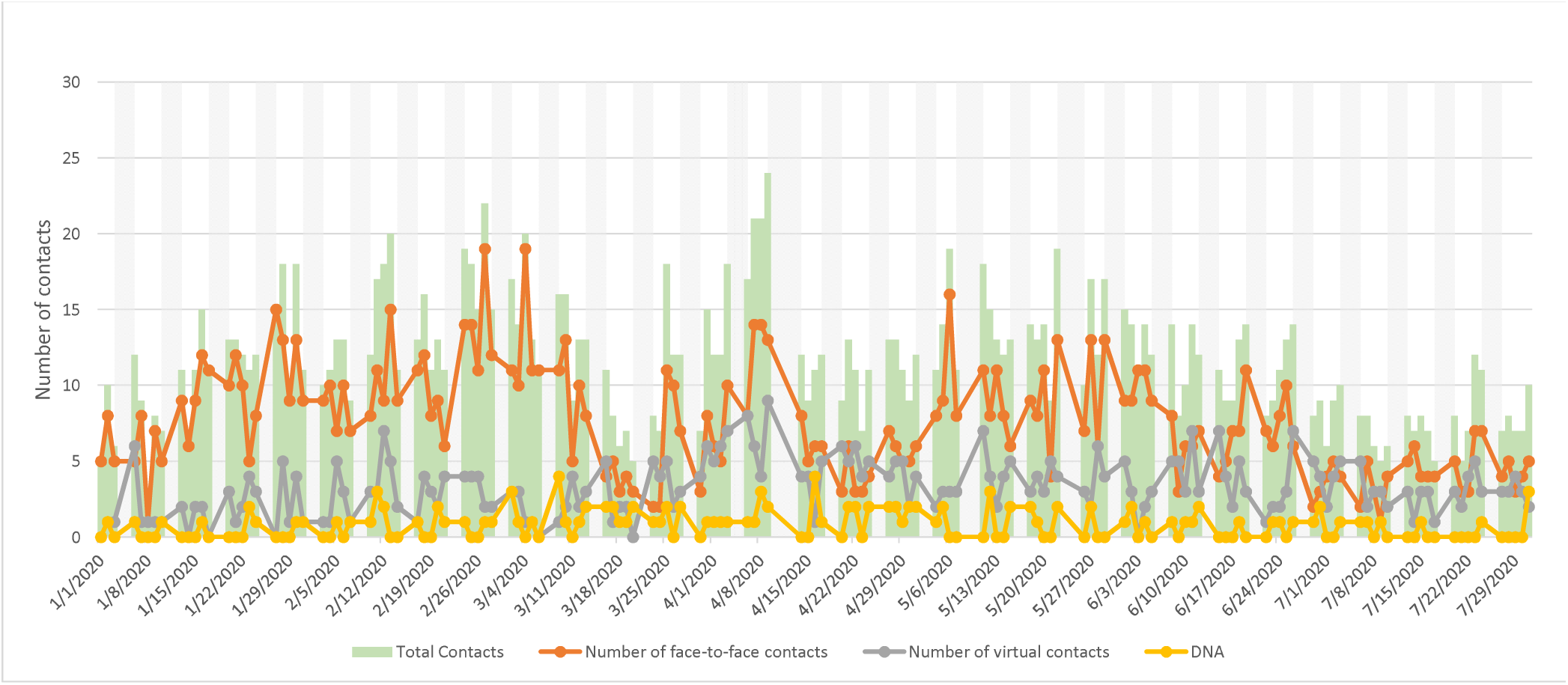
Home treatment / crisis services by type (daily; January - July 2020)

**Figure 13:**
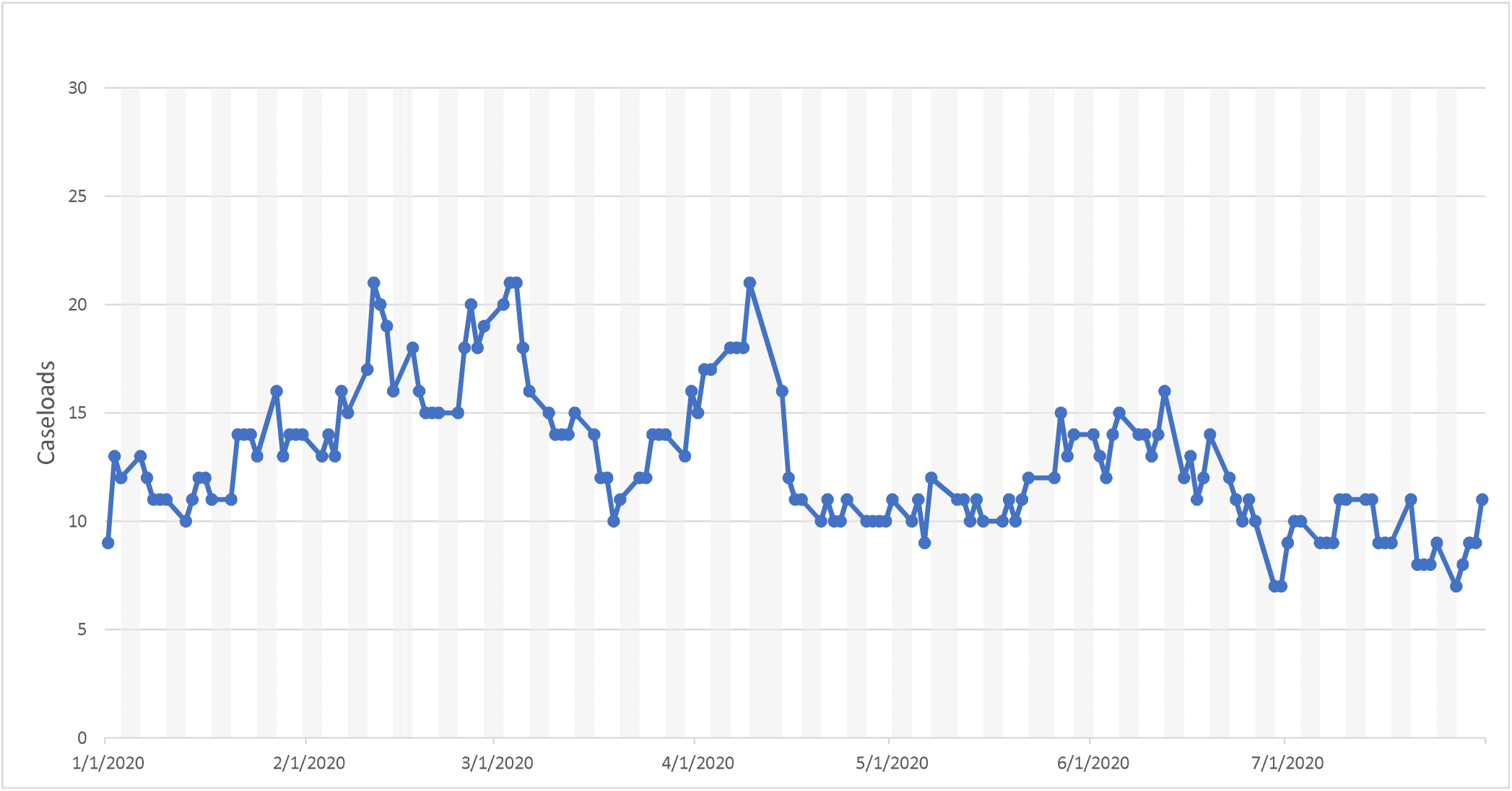
Home treatment / crisis services caseloads (daily; January-July 2020)

Liaison A&E daily contacts are displayed in Figure 14 and daily caseloads are displayed in Figure 15. Mean total daily contacts were reduced by 9%: a 13% reduction in face-to-face contacts and a 42% increase in virtual contacts. Mean daily caseloads were reduced by 15%.

**Figure 14:**
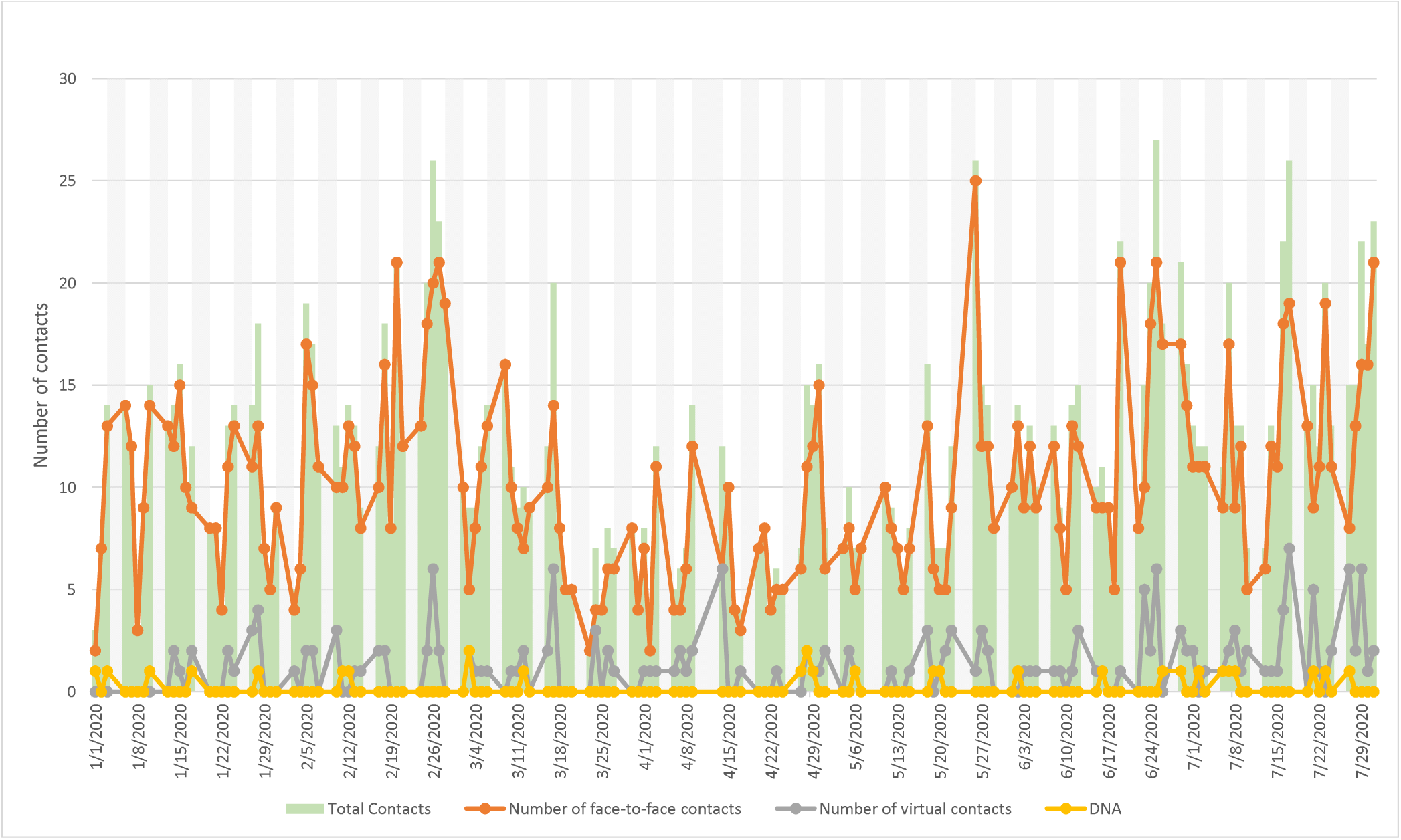
Liasion / A&E services contacts by type (daily; January - July 2020)

**Figure 15:**
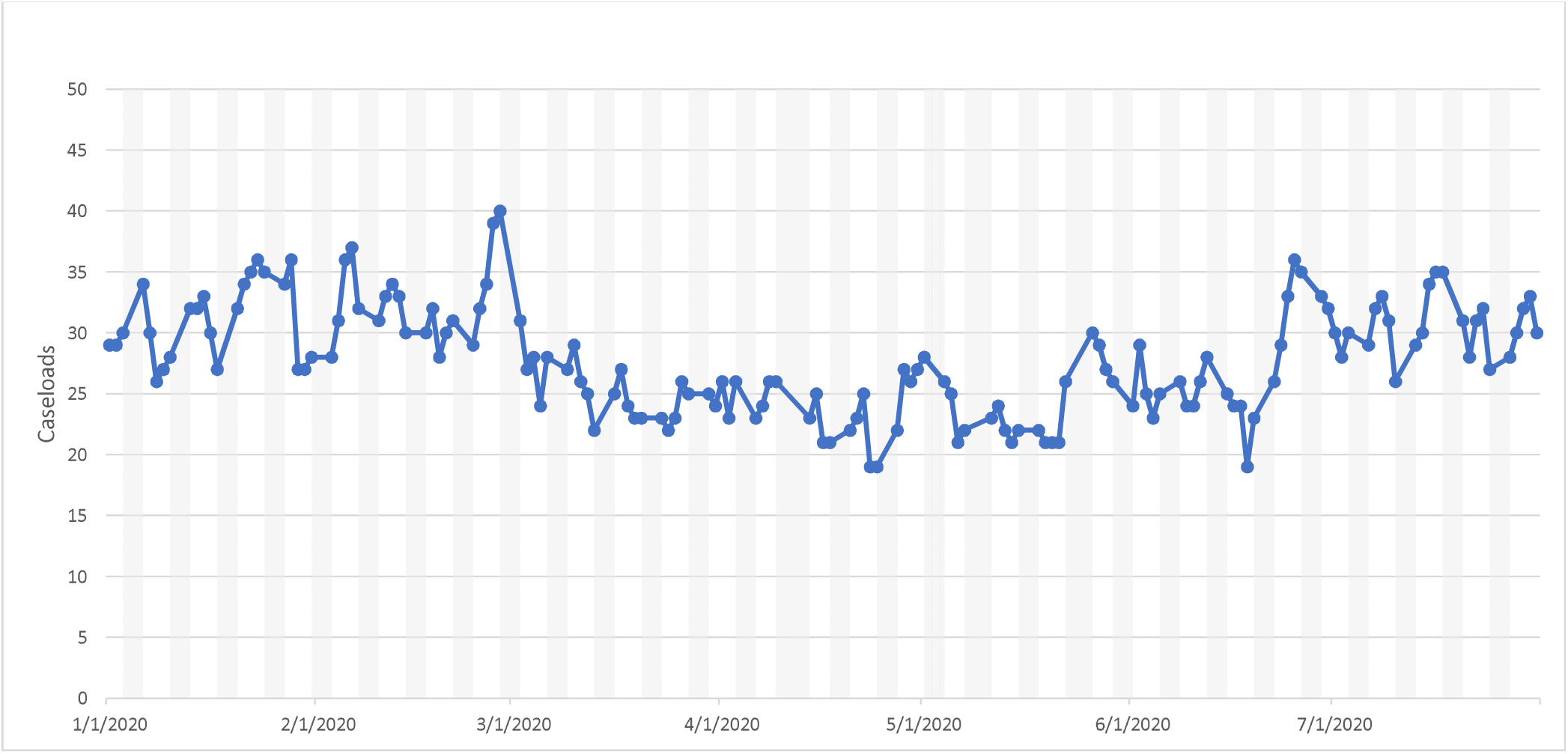
Liaison / A&E services caseloads (daily; January - July 2020)

MHOA daily contacts are displayed in Figure 16 and daily caseloads are displayed in Figure 17. Mean total daily contacts were reduced by 32%: a 47% reduction in face-to-face contacts and a 4% increase in virtual contacts. Mean daily caseloads reduced by 18%.

**Figure 16:**
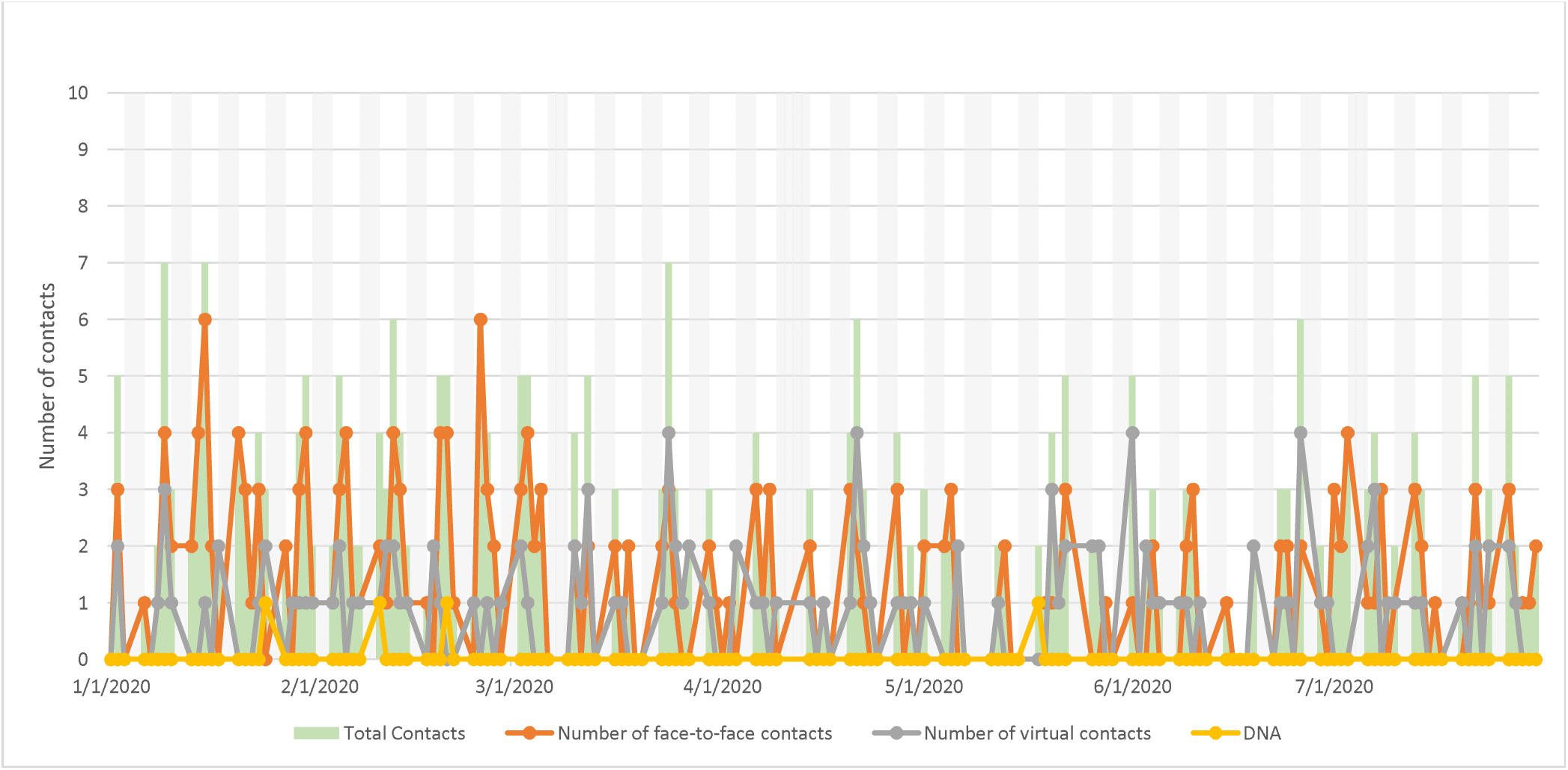
Older adult services contacts by type (daily; January - July 2020)

**Figure 17:**
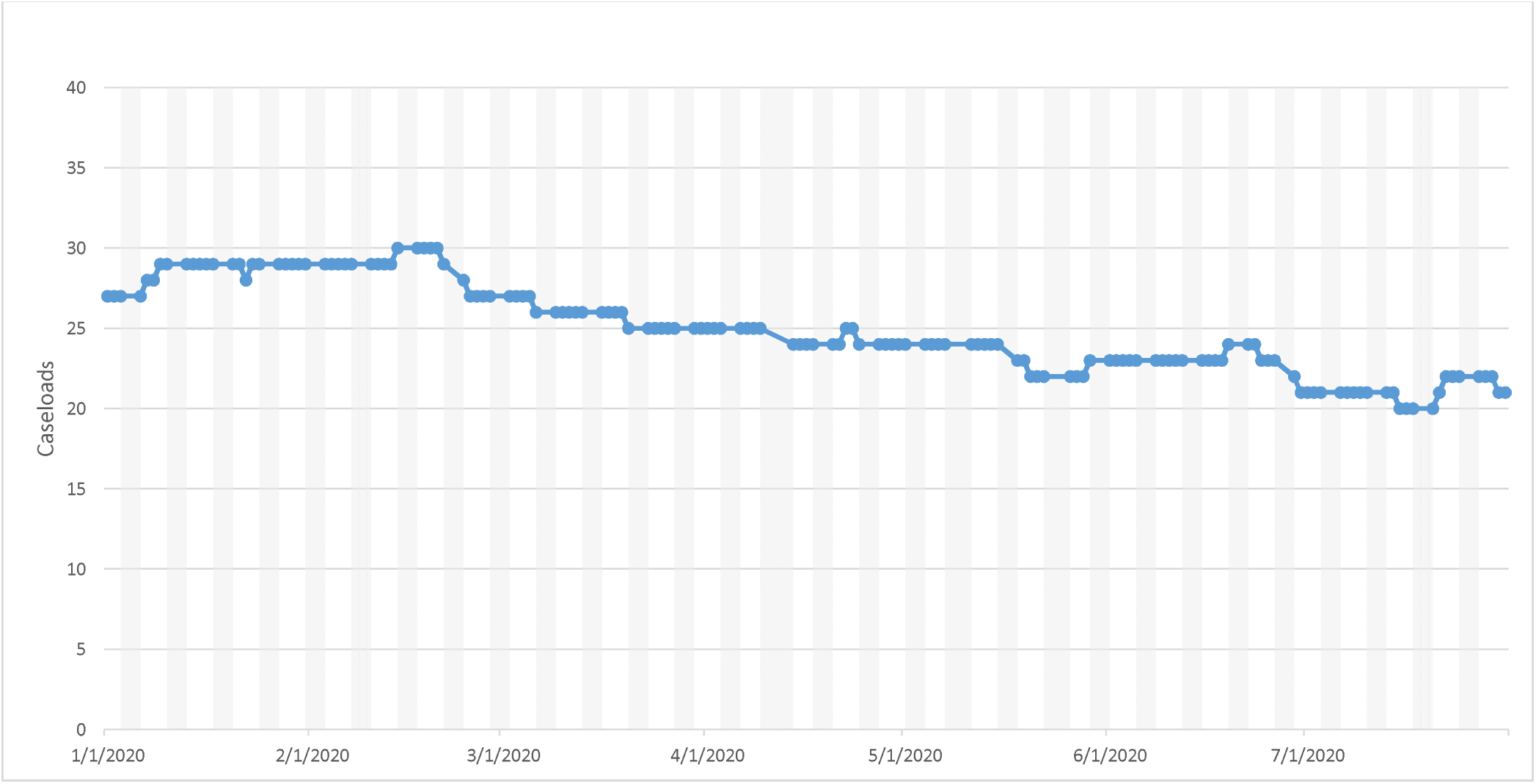
Older adults services caseloads (daily; January - July 2020)

## Discussion

Comparing periods before and after the 16^th^ of March 2020 in terms of the overall Trust activity for individuals with potential neurodevelopmental disorders, total caseload numbers remained relatively unchanged (a 6% decrease), although inpatient numbers were reduced more substantially (by 22%) because of discharges made in the early period of lockdown. Caseloads were reduced in all specific services, but most substantially in HTT (21%), MHOA (18%) and Liaison/A&E (15%), and were least changed in EIP (1%), AMHS (5%) and CAMHS (6%).

All specific services also recorded reductions in face-to-face and increases in virtual contacts. Total contact numbers declined most in MHOA (by 32%), followed by HTT (14%), Liaison/A&E (9%) and CAMHS (7%), and were increased in AMHS (by 6%) and EIP (17%). However, it should be borne in mind that contact numbers were relatively low in some services (MHOA in particular).

Mean daily deaths increased by 103% for the period after 16^th^ March compared to the daily mean for 2020 up to this date; however, the period from 16^th^ March to 31^st^ July included both the peak wave of increased mortality and a period with relatively few deaths. As described, the increase rose to 282% if the mid-March to mid-May period was considered specifically.

Considering limitations, it is important to bear in mind that the data are derived from a single site. Because complete data are being provided for that site with no hypothetical source population intended, calculation of confidence intervals was not felt to be appropriate for the descriptive data provided in this report; applicability to other mental healthcare providers cannot therefore be inferred and would need specific investigation. Profiles of services used by individuals with potential neurodevelopmental disorders and catchment morbidity are also likely to vary. It is also likely that we did not capture all of the services individuals with neurodevelopmental disorders are currently accessing for mental healthcare, as well as undiagnosed individuals; in addition, inclusion criteria defined a relatively broad group including learning disability and ADHD diagnoses. Classification of service types is inevitably approximate and likely to vary between mental health service providers. Finally, no attempt was made to adjust for temporal changes (e.g. seasonal effects) that might be observed in previous years’ data.

## Supporting information

STROBE checklist

## Data Availability

Data are available on request from the corresponding author.

## Funding

The research leading to these results has received support from a grant from King’s Together. EN and EM are supported by the ESRC-funded DETERMIND project. RS and MB are part-funded by the National Institute for Health Research (NIHR) Biomedical Research Centre at the South London and Maudsley NHS Foundation Trust and King’s College London; RS is additionally part-funded by: i) a Medical Research Council (MRC) Mental Health Data Pathfinder Award to King’s College London; ii) an NIHR Senior Investigator Award; iii) the National Institute for Health Research (NIHR) Applied Research Collaboration South London (NIHR ARC South London) at King’s College Hospital NHS Foundation Trust. The views expressed are those of the authors and not necessarily those of the NIHR or the Department of Health and Social Care.

## Notes

### Competing Interest Statement

RS declares research support in the past 36 months from Janssen, GSK and Takeda

### Funding Statement

The research leading to these results has received support from a grant from Kings Together. EN and EM are supported by the ESRC-funded DETERMIND project. RS and MB are part-funded by the National Institute for Health Research (NIHR) Biomedical Research Centre at the South London and Maudsley NHS Foundation Trust and Kings College London; RS is additionally part-funded by: i) a Medical Research Council (MRC) Mental Health Data Pathfinder Award to Kings College London; ii) an NIHR Senior Investigator Award; iii) the National Institute for Health Research (NIHR) Applied Research Collaboration South London (NIHR ARC South London) at Kings College Hospital NHS Foundation Trust. The views expressed are those of the authors and not necessarily those of the NIHR or the Department of Health and Social Care.

### Author Declarations

CRIS has received approval as a data source for secondary analyses (Oxford Research Ethics Committee C, reference 18/SC/0372)

### Summary of Updates

Following discussion with colleagues, we feel that 'potential neurodevelopmental disorders' better reflects the diagnostic groups being described than 'learning disabilities' in the original manuscript. Therefore we have changed this terminology and have added a sentence in the limitations section to acknowledge the mix of diagnoses being considered here.

